# Towards Personalized Breast Cancer Risk Management: A Thai Cohort Study on Polygenic Risk Scores

**DOI:** 10.1101/2024.07.28.24311135

**Authors:** Vorthunju Nakhonsri, Manop Pithukpakorn, Jakris Eu-ahsunthornwattana, Chumpol Ngamphiw, Rujipat Wasitthankasem, Alisa Wilantho, Pongsakorn Wangkumhang, Manon Boonbangyang, Sissades Tongsima

**Author notes:** Corresponding author: Sissades Tongsima.

## Abstract

Polygenic Risk Scores (PRS) are now playing an important role in predicting overall risk of breast cancer risk by means of adding contribution factors across independent genetic variants influencing the disease. However, PRS models may work better in some ethnic populations compared to others, thus requiring populaion-specific validation. This study evaluates the performance of 140 previously published PRS models in a Thai population, an underrepresented ethnic group. To rigorously evaluate the performance of 140 breast PRS models, we employed generalized linear models (GLM) combined with a robust evaluation strategy, including Five-fold cross validation and bootstrap analysis in which each model was tested across 1,000 bootstrap iterations to ensure the robustness of our findings and to identify models with consistently strong predictive ability. Among the 140 models evaluated, 38 demonstrated robust predictive ability, identified through > 163 bootstrap iterations (95% CI: 163.88). PGS004688 exhibited the highest performance, achieving an AUROC of 0.5930 (95% CI: 0.5903–0.5957) and a McFadden’s pseudo R^2^ of 0.0146 (95% CI: 0.0139–0.0153). Women in the 90^th^ percentile of PRS had a 1.83-fold increased risk of breast cancer compared to those within the 30^th^ to 70^th^ percentiles (95% CI: 1.04–3.18). This study highlights the importance of local validation for PRS models derived from diverse populations, demonstrating their potential for personalized breast cancer risk assessment. Model PGS004688, with its robust performance and significant risk stratification, warrants further investigation for clinical implementation in breast cancer screening and prevention strategies. Our findings emphasize the need for adapting and utilizing PRS in diverse populations to provide more accessible public health solutions.

## Introduction

Breast cancer is one of the main causes of death among women all over the world and is a multifactorial disease that depends on genetic and environmental factors [1]. Although some breast cancer cases are associated with strong penetrant mutations in genes such as *BRCA1* and *BRCA2*, most are associated with multiple low penetrant genetic variants [2]. This polygenic nature of breast cancer underscores the need for tools that can accurately assess an individual’s cumulative genetic predisposition. Polygenic risk scores (PRS), which aggregate the effects of these numerous common genetic variants, have emerged as a promising tool in this regard. PRS offer a quantitative measure of an individual’s genetic predisposition to breast cancer, potentially enabling more targeted screening and prevention strategies [3-4].

While the field of breast cancer PRS research is rapidly expanding, with over 140 models publicly available through repositories like the PGScatalog [5], a critical knowledge gap remains. The majority of these models were developed using data from Western populations, raising concerns about their accuracy and applicability across diverse ethnic groups [6]. Genetic and environmental variations between populations can significantly influence the performance of PRS, highlighting the urgent need for localized validation and adaptation of existing models. Furthermore, there is a lack of research on these models in Asian populations, especially in Southeast Asia. This absence in the development of PRS increases questions on the generalization of the current models to these groups. To fill this gap and facilitate the ability of PRS to accurately estimate breast cancer risk across ethnicities, regional studies, including this one involving a Thai cohort, are important [7-8]. This is crucial to ensure that PRS can effectively assess breast cancer risk in individuals from various backgrounds and ultimately contribute to more equitable and personalized healthcare.

This study aims to evaluate the performance of existing PRS models in a Thai cohort of breast cancer patients, contributing to a more comprehensive understanding of the generalizability and clinical utility of PRS for breast cancer risk assessment in diverse populations.

## Materials and Methods

### Study Population

This study utilized whole genome sequencing (WGS) data from 184 unrelated Thai women diagnosed with primary breast cancer who were treated at Siriraj Hospital. These data were obtained from previous studies, and the comprehensive case information was recently published [9]. To focus on the polygenic contribution to breast cancer risk, 38 patients harboring pathogenic or likely pathogenic (P/LP) variants in known breast cancer genes were excluded from the analysis (see Supplementary Table S1). The control group consisted of WGS data from 434 unrelated Thai individuals without cancer (Supplementary Table S2).

### Polygenic Risk Score Acquisition and Calculation

A total of 140 harmonized Polygenic Risk Scores (PRS) related to breast cancer (MONDO:0007254) were downloaded from the PGS Catalog on May 27^th^ 2024 [5]. To ensure compatibility with variant call format (VCF) data derived from WGS sequences, these scores were adapted using an in-house pipeline which involved normalizing the effect alleles to the GRCh38 reference genome using the BCFtools plugin `+fixref` [10] and adjusting the weight of the effect alleles aligned with the reference allele by multiplying them by -1. Each PRS was then calculated using the following formula:

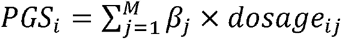

where *PGS*_*i*_ represents the polygenic score for the *i* ^th^ individual, β_*j*_ is the weight of the alternate allele at the locus *j*, and *dosage*_*ij*_ is the genotype dosage at that locus for the individual *i*.

### *Statistical* Analysis

To assess the robustness and generalizability of the PRS models, we employed a bootstrap analysis. In each of 1,000 bootstrap iterations, we randomly sampled 128 breast cancer cases and 128 controls to form a training set. Five-fold cross-validation was applied within this training set to identify the best-performing model for each iteration. Model performance was evaluated using McFadden’s Pseudo R^2^ and the log-likelihood ratio p-value to assess goodness of fit [11]. The Area Under the Receiver Operating Characteristic Curve (AUROC) was calculated to evaluate the discriminatory ability of each model within an independent test set comprising 56 breast cancer patients and 306 controls. Models were ranked based on the frequency of achieving a statistically significant log-likelihood ratio p-value (<0.05) across the 1,000 bootstrap iterations. The final best-performing model was selected based on the average McFadden’s Pseudo R^2^ and AUROC values across all iterations. All statistical analyses were performed using the R programming environment [12-15].

### Language and Computational Tools

This manuscript was refined using the language model ChatGPT for linguistic and structural improvement of the text. [16]

## Results

### Performance of Polygenic Risk Scores in Predicting Breast Cancer Risk

A comprehensive bootstrap analysis was conducted on 140 PRS models, using 1,000 iterations to evaluate their ability to predict breast cancer status. Results indicated that, on average, each model demonstrated a statistically significant association with breast cancer status in 142.76 out of the 1000 bootstrap iterations (95% confidence interval: 122.57–163.88). A detailed breakdown of the performance of each model across the bootstrap iterations is provided in Supplementary Table S3, and a visual representation of the distribution of significant associations is shown in Figure 1A.

**Figure 1:**
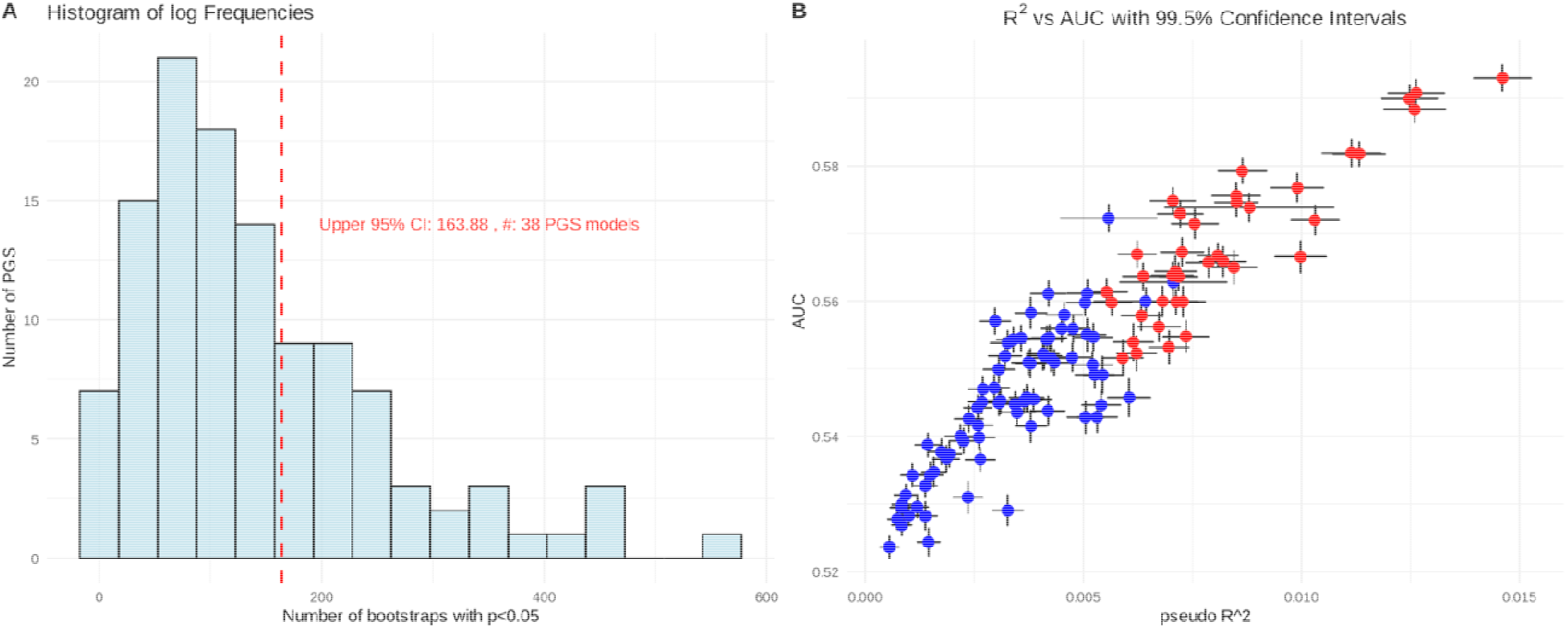
Bootstrap Performance of Polygenic Risk Scores for Breast Cancer Prediction. **(A) Distribution of Significant Associations:** Histogram displaying the number of bootstrap iterations (out of 1,000) in which each of the 140 PRS models achieved a statistically significant association with breast cancer status (p-value < 0.05). The red dashed line indicates the upper 95% confidence interval (163.88 iterations), highlighting models with frequent significant results. **(B) Predictive Performance and Consistency:** Scatter plot illustrating the relationship between McFadden’s Pseudo R^2^ and Area Under the Receiver Operating Characteristic Curve (AUROC) for each PRS model. Red dots represent models achieving significance in over 95% of bootstrap iterations, indicating high predictive consistency.

To further evaluate the performance, we plotted McFadden’s Pseudo R^2^ against AUROC for each model, including 95% confidence intervals (Figure 2A). This analysis identified PGS004688 as the top-performing model, demonstrating the highest average AUROC (0.5930; 95% CI: 0.5903–0.5957) and Pseudo R^2^ (0.0146; 95% CI: 0.0139–0.0153). Figure 2B provides a detailed visualization of the 1,000 bootstrap iterations for PGS004688, with a green square highlighting the mean ± 95% CI of both train and test AUROC values.

**Figure 2:**
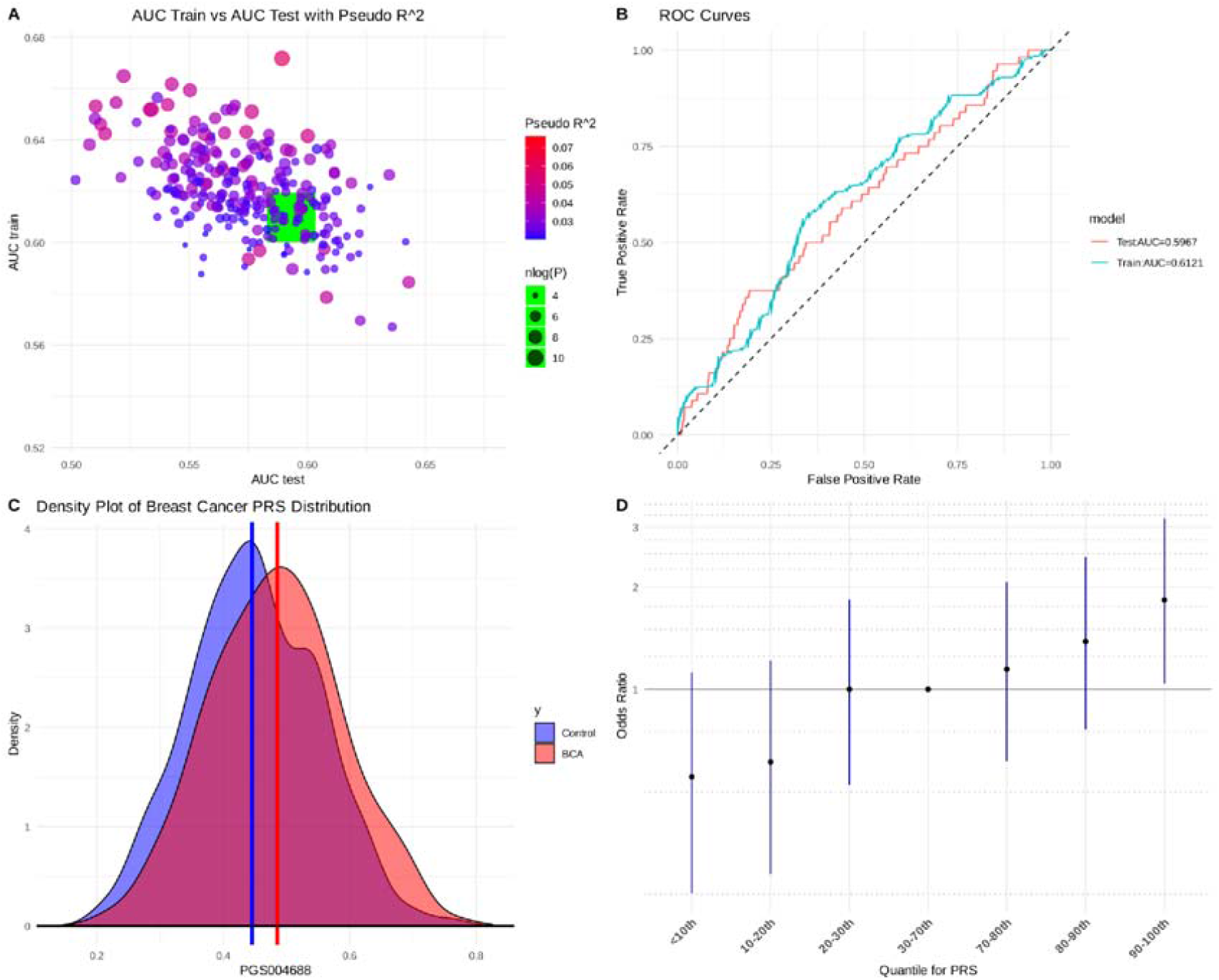
PGS004688: Predictive Accuracy and Consistency. **(A)** Scatter plot depicting McFadden’s Pseudo R^2^ versus AUROC for each PRS model. PGS004688 is highlighted, with a green square indicating the mean and 95% confidence interval of AUROC values. **(B)** ROC curves for PGS004688, comparing performance in the training (green) and testing (red) datasets to demonstrate model consistency. **(C)** Density plot illustrating the distribution of standardized PGS004688 scores in breast cancer patients (red) and controls (blue), with median scores indicated. **(D)** Forest plot displaying odds ratios for breast cancer risk at different PGS004688 quantiles. Notably, individuals with scores above the 90th percentile exhibit a significantly elevated risk (odds ratio = 1.83; 95% CI: 1.04–3.18), highlighting the potential clinical utility of PGS004688 for risk stratification.

## Discussion

This study underlies the crucial need for population-specific validation of Polygenic Risk Scores (PRS), for accurate breast cancer risk management. Our findings demonstrate that PRS performance can vary significantly across different ethnicities due to variations in genetic diversity and allele frequencies [6]. This discrepancy is particularly evident when comparing European ancestry populations to more genetically diverse populations. While resources like the PGScatalog, containing over 4,000 PRS from over 600 studies, are invaluable, our study highlights the challenges of applying models developed in one population to another.

To address this, we adapted 140 breast cancer-related PRS for use with our Thai cohort. We employed rigorous cross-validation and bootstrap methods to ensure robust model generalization. Notably, we identified PGS004688 as the most effective PRS for predicting breast cancer risk in Thai women. Interestingly, despite being originally developed using GWAS data from a predominantly European cohort [17-18], PGS004688 outperformed models specifically developed for East Asian populations [19-20], This finding underscores the complexity of PRS transferability and the need for population-specific validation. While PGS004688 demonstrated superior performance in our Thai cohort, its effectiveness was lower than its reported performance in European ancestry cohorts (AUROC = 0.665) [18]. This disparity emphasizes the need for continued research and validation of PRS in diverse populations. Further investigation in larger Thai cohorts is crucial to confirm the clinical utility of Ensuring the clinical utility of PGS004688 and ensure its reliability for breast cancer risk assessment in Thailand.

## Conclusion

This study highlights the critical need for population-specific validation of Polygenic Risk Score (PRS) for accurate breast cancer risk assessment. Our findings demonstrate that PRS performance can vary significantly across different ethnicities due to variations in genetic diversity and allele frequencies. While resources like the PGScatalog are invaluable, our study reflects the challenges of applying models developed in one population to another. We identified PGS004688 as the most effective PRS for predicting bresat cancer risk in Thai women, outperforming models specifically developed for Eas Asian populations. This finding reveals the complexity of PRS transferability and the need for continued research and validation in diverse populations. Further investigation in larger Thai cohorts is imperative to confirm the clinical utility of PGS004688 and ensure its reliability for breast cancer risk assessment in Thailand.

## Data Availability

All data produced in the present study are available upon reasonable request to the authors

